# A mixed methods process evaluation of a peer coaching intervention to improve the implementation of preventive tasks by occupational physicians

**DOI:** 10.1101/2024.11.07.24316899

**Authors:** S. Orhan Pees, S.H. van Oostrom, F.G. Schaafsma, K.I. Proper

## Abstract

**Objectives:** This study aimed to evaluate the process of implementation of a peer coaching intervention program for occupational physicians (OPs) to improve the execution of preventive tasks. Specifically, the evaluation seeks to: (1) describe the reach and uptake of the intervention program; (2) determine the extent to which the program was implemented as intended; (3) provide insights into experiences of OPs, and (4) identify factors influencing the implementation.

**Methods:** This study employed a mixed-methods approach to assess seven main process indicators: acceptability, adoption, appropriateness, feasibility, fidelity, penetration and sustainability. Data were collected between March and June 2024 by means of an online questionnaire (N=98), and 17 semi-structured interviews with group coordinators and OPs.

**Results:** 20 out of 21 groups allocated to the intervention program participated in the intervention and 98 out of 115 participants (85%) filled in the questionnaire. Three-quarters of the participants completed the entire program. 96% of the OPs successfully discussed barriers to the execution of preventive tasks, and 83% were able to formulate strategies for these barriers. Most participants managed to implement their formulated goals in practice. When they were unable to do so, time constraints and resistance from employers and their occupational health services often played a role. OPs valued the program’s structure, interaction with colleagues, and the increased awareness it generated.

**Discussion and conclusion:** The peer coaching group program was well-implemented and positively evaluated by OPs. The program can be improved by allocating more time to it, for instance by integrating it into the educational curriculum, and by paying more attention to the specific working conditions of OPs, such as the different sectors in which they are employed.

**Trial registration:** ISRCTN registry; ISRCTN15394765. Registered on 27 June 2023.

## Introduction

Work-related health complaints are a major public and occupational health issue. In the European Union alone, 10.3% of all workers reported having work-related health problems, with 18.6% of these due to mental health problems [1]. Work-related health problems are estimated to account for a loss of approximately 4-6% Gross Domestic Product (GDP) [2]. These high numbers of people facing work-related health-complaints and economic consequences emphasize the importance of prevention, such as assessment and management of occupational hazards, health surveillance and health promotion activities [3, 4]. However, the integration of prevention and implementation of preventive tasks in practice is often hampering. Factors playing a role in this are, among others, budgetary constraints, lack of awareness about the value of prevention and return of investment among management, insufficient multidisciplinary collaboration, and lack of knowledge and skills of OPs [5–7]. This is the case in the Netherlands as well, where preventive tasks are a legally prescribed and essential part of the work of occupational physicians (OPs). However, due to barriers experienced, OPs’ work is predominantly centred on absenteeism guidance and reintegration tasks.

In order to support OPs in their preventive role and to promote the execution of preventive tasks targeting work-related mental health problems, a peer coaching intervention program was developed following the Implementation Mapping protocol [8]. The program was based on identified barriers and facilitators for the execution of preventive tasks and consisted of three peer coaching meetings, in which prevention of work-related (mental) health problems was the main topic [9]. The primary objective of the peer coaching intervention program was to create a collaborative learning environment, in which the exchange of knowledge, experiences, and ideas with regard to prevention between OPs was central. During the meetings, OPs addressed perceived barriers with regard to the execution of preventive tasks, discussed possible strategies to overcome these barriers with their colleagues, focused on sharing best practices, and developed a personalized action plan to increase the implementation of preventive tasks in practice, using input from colleagues [9].

The IM-PROmPt (Implementation of PReventive tasks by Occupational Physicians) study was designed to develop and evaluate the peer coaching intervention program. The IM-PROmPt study is a two-armed cluster randomized controlled trial (c-RCT), comparing the developed intervention program directed to the implementation of preventive tasks for OPs with usual peer group meetings. To increase insights in the implementation of the program and help to explain its potential effectiveness, a process evaluation was conducted alongside this randomized controlled trial [10]. Process evaluations are essential for understanding why an intervention does or does not produce the desired effects, which factors or mechanisms influence these outcomes, whether the intervention was implemented as intended and whether participants received adequate exposure to it [11–13]. This is especially of importance in complex (group) interventions [14] and complex multisite trials where one intervention could have been implemented in different ways [15], as is the case in our study where the intervention was implemented into existing peer groups. Moreover, process evaluations provide valuable insights into the experiences of participants and users, offering a better understanding of how the intervention is perceived and utilized in real-world settings [13]. These insights are important for identifying areas for improvement and for adapting the intervention to better meet the needs of its target population.

This process evaluation served four aims, of which the first was to describe the reach and uptake of the peer coaching intervention program. The second aim was to examine the extent to which intervention program was implemented as intended. The third aim was to identify barriers and facilitators that may have impacted the implementation. Finally, this evaluation was aimed at examining the experiences and perspectives of OPs, perceived fit and benefits of the intervention program.

## Methods

### Study design

The peer coaching intervention program was implemented between September 2023 and May 2024. Data for this process evaluation were collected from participants randomized to the intervention condition between March and June 2024 by means of an online questionnaire and semi-structured interviews. In this mixed-methods process evaluation, the implementation outcomes framework as outlined by Proctor was utilized, which includes the dimensions of acceptability, adoption, appropriateness, costs, feasibility, fidelity, penetration, and sustainability [16]. This framework helps to understand the implementation of interventions across various settings. Specifically, our intervention program was applied within existing peer coaching groups, making it crucial to assess how well it integrated into these pre-established structures. These implementation outcomes also align well with the development of the program, which was guided by the principles of implementation mapping [8, 9]. An explanation of all indicators and how they have been assessed can be found in Table 1.

**Table 1:**
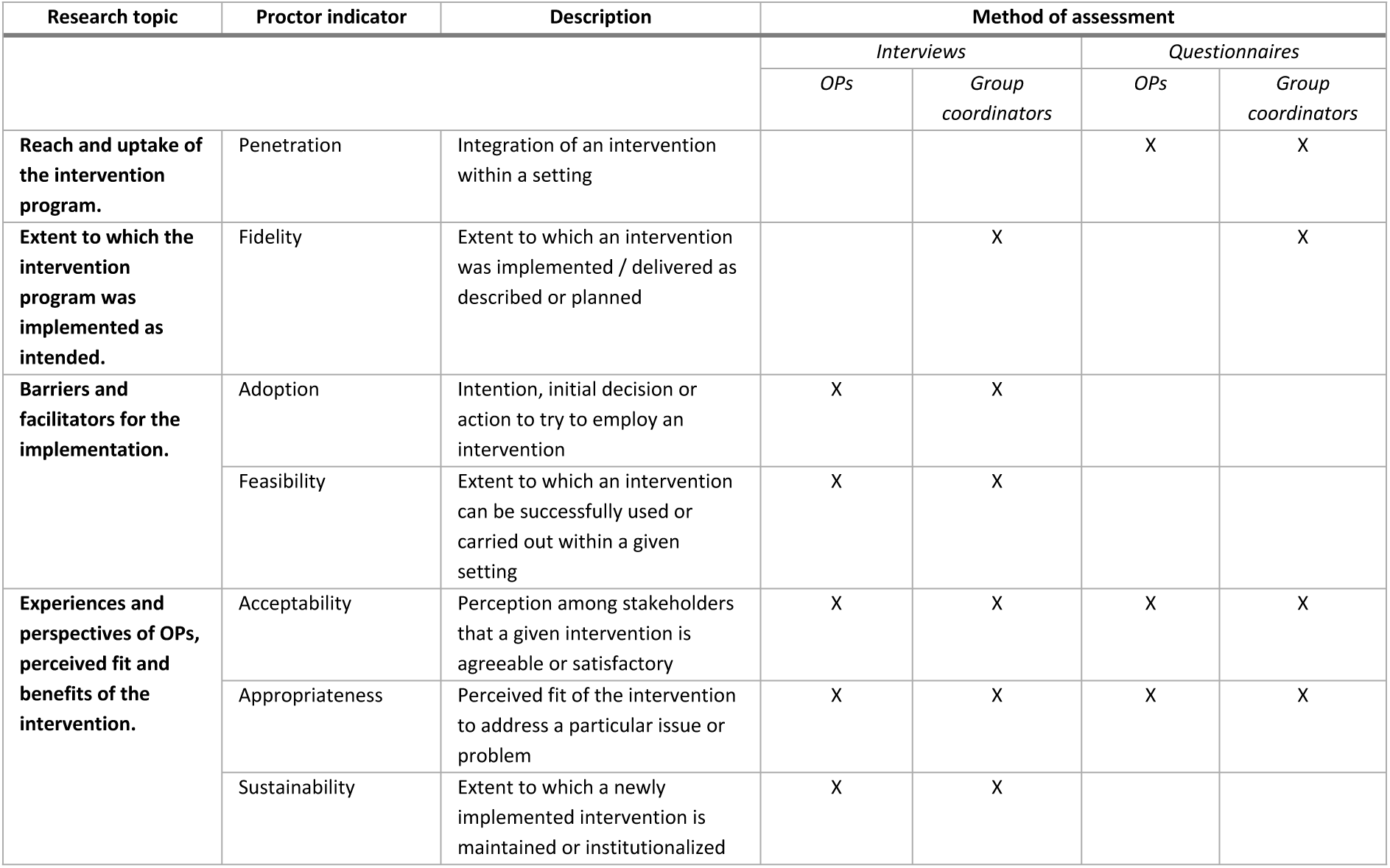
Process indicators and method of assessment.

This study was performed in accordance with the ethical guidelines and regulations laid down in the Declaration of Helsinki. The Medical Ethics Review Committee of the Academic Medical Center Amsterdam, the Netherlands, confirmed that the Medical Research Involving Human Subjects Act (WMO) does not apply to the study and that an official approval of this study was not required (reference number W22_415 # 22.490). Written informed consent was obtained from all participants via an online form prior to participating in this study.

### Peer coaching group intervention to implement preventive tasks

The intervention program to support OPs in their preventive role has been developed based on barriers and facilitators described in literature regarding the implementation of preventive tasks, input from stakeholders, and evidence-based strategies from literature [9]. The program incorporated strategies and methods from literature, including ongoing training, active learning, goal setting, peer education and discussion [17, 18].

Involved stakeholders and OPs expressed a desire not to establish new structures but to utilize existing structures and resources. In the Netherlands, OPs are required to attend peer group meetings as part of the Continuing Medical Education (CME) system, in order to receive accreditation points and maintain their licensure. Therefore, the program was implemented into these existing peer groups, which means that participants often already know each other. Group sizes ranged from 4 to 12 OPs, and one or two OPs were appointed as group coordinators.

The intervention program consisted of three group meetings over six months with a total duration of approximately five working hours. Coordinators of each group received an interactive two-hour training and associated instructions from the research team before the groups started organizing their meetings. Each session had its own theme with corresponding assignments before and during the meeting. During the first meeting, OPs discussed barriers experienced regarding prevention and individually set goals related to executing preventive tasks, which they worked on between the intervention meetings. The second meeting focused on the Prevention Cycle, a tool developed for Dutch OPs to help transform information from sick leave guidance into preventive actions (e.g. targeted at employees in the organisation), and discussing cases from practice. The third meeting focused on sustaining the topic of prevention in their work and how to keep prevention on the agenda of their peer group. Participating OPs reflected on their set goals throughout the course of the meetings, using insights and experiences from fellow OPs. Peer groups were equipped with educational materials regarding prevention and strategies to integrate preventive tasks into their daily practice. Special attention was given to prevention of work-related mental health problems in the materials and throughout the intervention program.

### Study population and recruitment

Participants in this study were OPs working in the Netherlands. Recruitment took place between May 10 and July 10, 2023, via email invitations. Group coordinators were invited to participate with their groups, after which individual participants within each group received the online application and informed consent form. OPs were excluded from the study if they (1) had an upcoming retirement or long-term leave (e.g., maternity leave) planned during the study follow-up, or (2) worked fewer than 16 hours per week in OP practice (with hours dedicated to research, education, or policy excluded). The baseline study population consisted of 227 OPs, of whom 124 were assigned to the intervention condition and subsequently answered questions regarding the implementation process immediately after completing the intervention program, between March 8 and June 21, 2024. Further information on recruitment and inclusion criteria can be found in the study protocol [9].

### Data collection

The questionnaire focused on the constructs penetration, fidelity, acceptability and appropriateness [16, 19]. Penetration (e.g. dose received) involved reporting the number of attended meetings (0-3) and reasons for non-attendance. Acceptability was evaluated through rating different statements about participants’ experience with the intervention program on a five-point Likert scale. For example, “*The intervention program aligned well with my aim to dedicate more time to the execution of preventive tasks*”. Additionally, participants rated various aspects of the program on a scale from 1 (worst) to 10 (best) (e.g. “*Which grade would you give to the intervention guide with tips and assignments*?”). Moreover, participants were asked to highlight what they liked most about the program and suggest ideas for improvement. Group coordinators received additional questions designed to evaluate the constructs fidelity, to assess the extent to which groups and group coordinators succeeded in adhering to the program as described in the training and additional materials. These fidelity-related questions were measured on a five-point Likert scale (e.g. “*Have you managed to discuss the wishes and challenges regarding preventive tasks with the members in your ICT group*”).

Coordinators of each peer group (N=21) allocated to the intervention condition received additional questions regarding their experiences with organizing and facilitating the intervention meetings. In the questionnaires, participants could indicate whether they were also willing to participate in an online interview. Individual semi-structured interviews were held between March and May 2024 with 9 group coordinators and 8 participating OPs. The interviews aimed to assess all process indicators. For a thorough assessment of the constructs acceptability, appropriateness and fidelity, five determinants from the Measurement Instrument for Determinants of Innovations (MIDI) were included in the interview guide: procedural clarity; completeness; complexity; compatibility; and relevance [20].

### Data analysis

The online interviews were audio recorded and transcribed verbatim. The transcripts were analysed by means of thematic coding by two independent researchers, using MAXQDA software. Because of the use of clearly defined frameworks for the development of the interview guide, transcripts were coded deductively. Author SOP and an independent research assistant individually analysed a subset of transcripts, followed by discussion to reach consensus on interpretations and codes. Subsequently, SOP analysed the remaining transcripts, and the research assistant reviewed the codes. The codes were collaboratively examined to identify patterns and themes. The final list of codes and themes were discussed within the research team. Descriptive statistics were used to analyse the closed survey questions regarding fidelity and acceptability, making use of statistical software Stata/SE version 18.0. Differences in outcomes were tested between the high compliant group (i.e. attending all intervention meetings) and the low compliant group. The open-ended questions in the questionnaire focused on the set goals and the extent to which they were achieved and these were analysed using MAXQDA software.

## Results

### Reach and uptake

#### Penetration

The Dutch Professional Association for Occupational Physicians (NVAB) sent out e-mail invitations to all group coordinators in May 2023, reaching approximately 360 groups. Of these, 41 groups registered for participation via an online form, after which individual OPs from these groups received an information letter with a link to an online consent form to share with the individual OPs in their group. Group sizes ranged from four to twelve members. Four out of 45 groups did apply for participation but did not meet the required number of three participating OPs in their group. A total of 21 out of 41 participating peer groups was allocated to the intervention condition, but one group later withdrew from the study due to other priorities and the desire to participate in the program at a later time. A total of 115 participants was included in the intervention condition, of which 98 participants (85%) in 20 groups filled in the questionnaire (23 group coordinators and 75 participating OPs). Participants worked on average 36 hours per week. With regard to size of the organizations, approximately 30% of participants was contracted by small and medium-sized enterprises (SME), while the rest worked for larger organizations. A flow chart of the study can be found in Figure 1.

**Figure 1:**
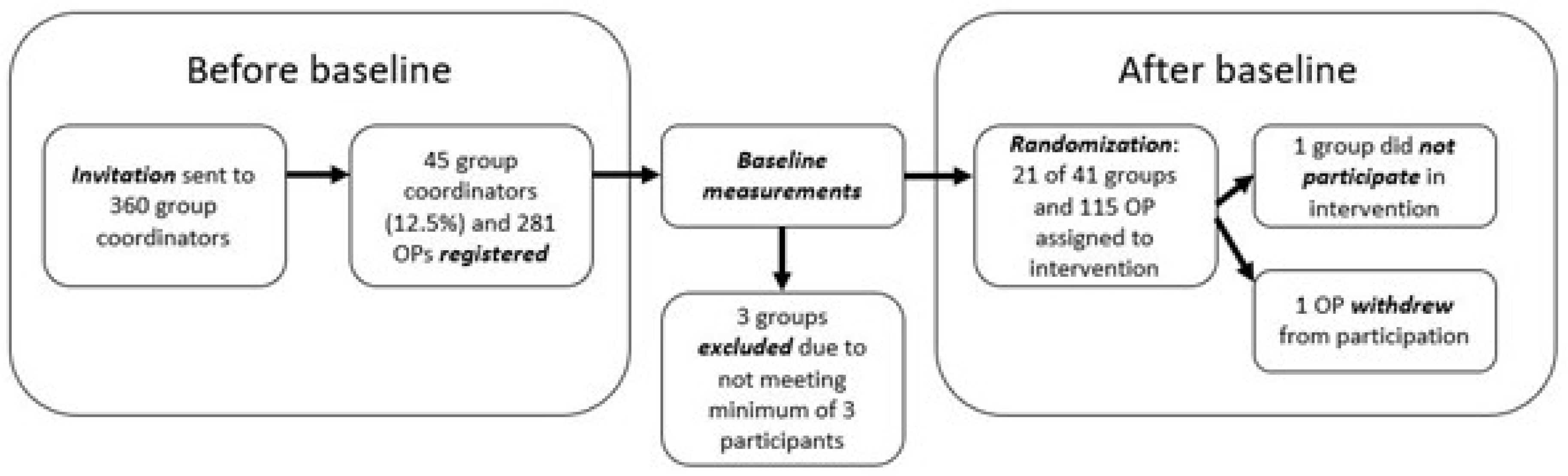
Flowchart of recruitment and inclusion of participants

Approximately nine months after organizing the online training for group coordinators, 17 out of 20 groups had organized all three meetings. Three groups had not yet organized the third meeting, due to organizational reasons (e.g. time) or too many absences of OPs. The first and second meetings were attended by approximately 90% of OPs, while the third meeting was attended by 80%. In total, 70 participants attended the full program and 28 participants attended partially. Reasons for non-attendance included illness, vacation, other commitments and obligations, or personal circumstances. No statistical significant differences were found in outcomes between the high compliant group and the low compliant group.

### Implementation as intended

#### Fidelity

To assess the extent to which the intervention program was delivered and followed as intended, group coordinators were asked to indicate if they had succeeded in different aspects of the program. The vast majority of group coordinators indicated that they mostly or fully succeeded in discussing challenges regarding prevention with their group (96%) and in discussing possible solutions to overcome these challenges (83%). Group coordinators were also asked to indicate the extent to which they succeeded in engaging their group and actively working on the theme of prevention and preventive tasks. In this regard, 70% (N=16) indicated that they had fully or mostly succeeded, 26% (N=6) that they had partly succeeded and partly not, and 4% (N=1) that they had mostly not succeeded (see Figure 2).

**Figure 2:**
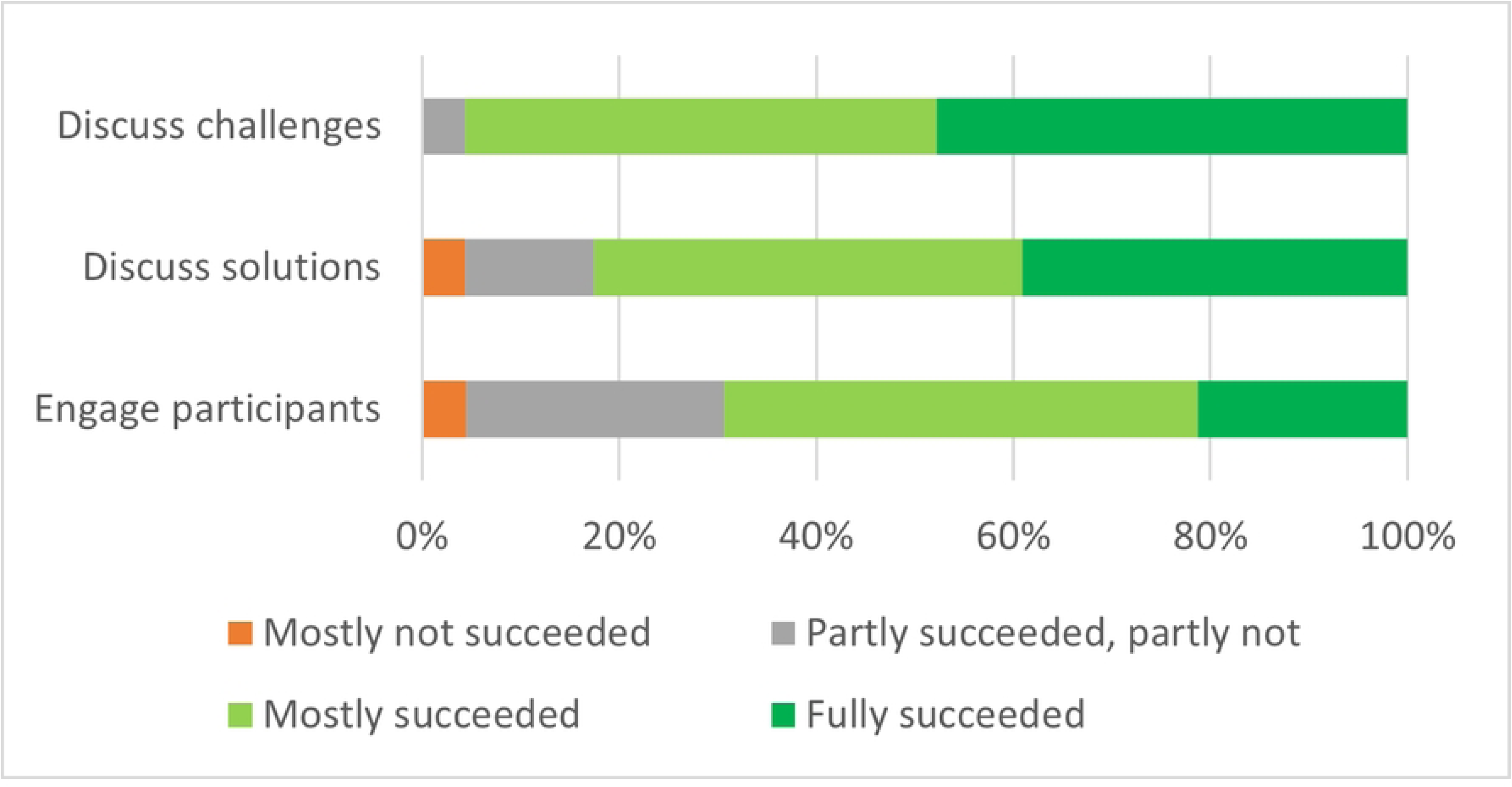
Statements on fidelity, scored by group coordinators (N=23)

One of the key elements of the program was to formulate (and subsequently implement) an action plan with attainable goals to achieve during the course of the program. A large number of participants (N=30) aimed to discuss prevention with the employer, for example by blocking time in their own schedules: *“That has been a goal for me: to set my own agenda, at least for one half-day. I am still succeeding at that. […] I hope to expand this to other days as well, because prevention is not specifically focused on one afternoon. I need to try to keep those other days free too, so that I can work on things in between.” [Interview 10; group coordinator].* Other goals frequently mentioned in the questionnaire related to organizing workplace visits (N=14), applying prevention in absence consultation (N=12), and organizing Periodic Medical Examination and Periodic Occupational Health Examinations (N=11). Several group coordinators explained during the interviews that they had not set their own goals but had primarily taken the role of process facilitator. They focused more on leading the program effectively and supporting their colleagues in developing and achieving their goals, rather than actively working on an action plan themselves.

In the program and associated materials, specific attention was given to preventing work-related mental health issues such as stress complaints. However, it turned out that most participants did not explicitly focus on these issues when setting their goals and action plans. Instead, they opted for more general and overarching actions such as visiting the workplace to assess risks or scheduling more meetings with clients and employers. A small number of participants indicated in the questionnaire and interviews that they were unable to set goals for themselves. Contextual barriers often played a role in this, as was explained during one interview: *“It was not really possible to set a goal, at least not in terms of concrete actions we could take. Of course, we had ideas on how to structure a preventive medical examination, but how to actually implement it? We couldn’t figure that out. We neatly addressed all those questions… We had everything in order, but every time we ended up with the same issue: but how exactly?” [Interview 06; group coordinator]*

In the questionnaire, 39 OPs explicitly stated that they (partially) succeeded in achieving their goals, while 26 OPs stated that they were not able to achieve their goals. Resistance from employers often played a role in this, especially in smaller organizations. Moreover, one OP working for Occupational Health and Safety services (OHS) experienced limited influence over their own client base and the content of the contracts regarding preventive tasks, leading to a mismatch in expectations between the OP and companies they served. Additionally, due to a lack of time, it was sometimes not possible to achieve their goal within the duration of the study, but plans were made to continue working on it, and sometimes follow-up steps were already scheduled: *“Well, yes, my goal was to work more with the Works Council and other parties. And with the Works Council, the first bridge has been built, but I notice that there is still a long way to go because the council is not really focused on prevention. […] It definitely deserves follow-up; it’s not something that can be accomplished quickly.” [Interview 09; group coordinator]*.

### Barriers and facilitators for the implementation

#### Adoption

During the interviews, participants were asked for their reason to participate and their expectations beforehand. It appeared that most participants participated in the program because they find prevention important but feel they do not spend enough time and attention to it in their daily work as an OP. Additionally, curiosity also appeared to play a role, and participants were eager to see what they could learn from their colleagues: “*An important reason for signing up was to see what else could be learned, in addition to curiosity about how it would be approached. And also to engage in discussions with other colleagues […]: What do we not know about each other yet? What has never been brought to the surface?" [Interview 02; group coordinator]*

Although almost all group coordinators indicated that all participants in their group had agreed to participate, it became apparent that some participants’ attention and interest decreased over time. Several coordinators mentioned that it was challenging to keep everyone actively involved: *"It was difficult to keep the whole group engaged. There was a bit of variation. I said beforehand: ‘Guys, if we’re going to participate, you need to show commitment. So if you start, I think you should attend all the meetings, you need to be there.’ But not all participants managed to do that." [Interview 12; group coordinator]*

#### Feasibility

Most participants mentioned that the proposed frequency of meetings worked well, because it provided a reminder to focus on preventive tasks or aligned well with their planned meetings. However, a minority of participants felt that the intervals could be longer, allowing more time and opportunities between meetings to address preventive tasks. Although it was not specifically asked, several participants mentioned that the time allocated for the meetings (2 hours, 1.5 hours and 1 hour, respectively) was not sufficient and that they needed more time: *“I believe it took more time than expected in the beginning. I remember the first meeting was supposed to be an hour or 90 minutes or something like that. I think we spent more time on it, especially the first time.” [Interview 04; group coordinator]*

As previously explained under the section *Fidelity*, the majority of participants successfully formulated goals for themselves. Most participants were also able to (partially) achieve these goals within the program’s timeframe, although additional time was sometimes needed to complete the action plan. Time was frequently mentioned as a reason when participants were unable to achieve their goals. Other common barriers included personal factors such as illness and employers’ low priority for prevention which involved the OP.

### Experiences and perspectives of OPs

#### Acceptability

On a scale from 1 (worst) to 10 (best), the three meetings were scored with an average of 7.6, 7.6 and 7.4, respectively. The developed materials for participants was rated a 7.4, while the instructions and training for group coordinators were both rated a 7.9. Group coordinators rated themselves a 7.3 for their role as group facilitators, while participants rated the coordinators’ role an 8.2. The full program was rated a 7.5. A complete list of the given ratings can be found in Table 2.

**Table 2:**
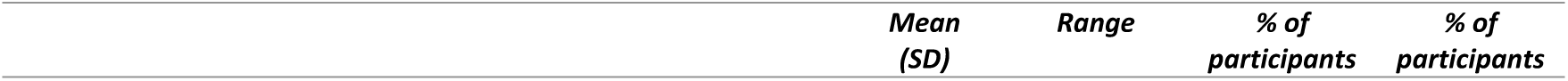
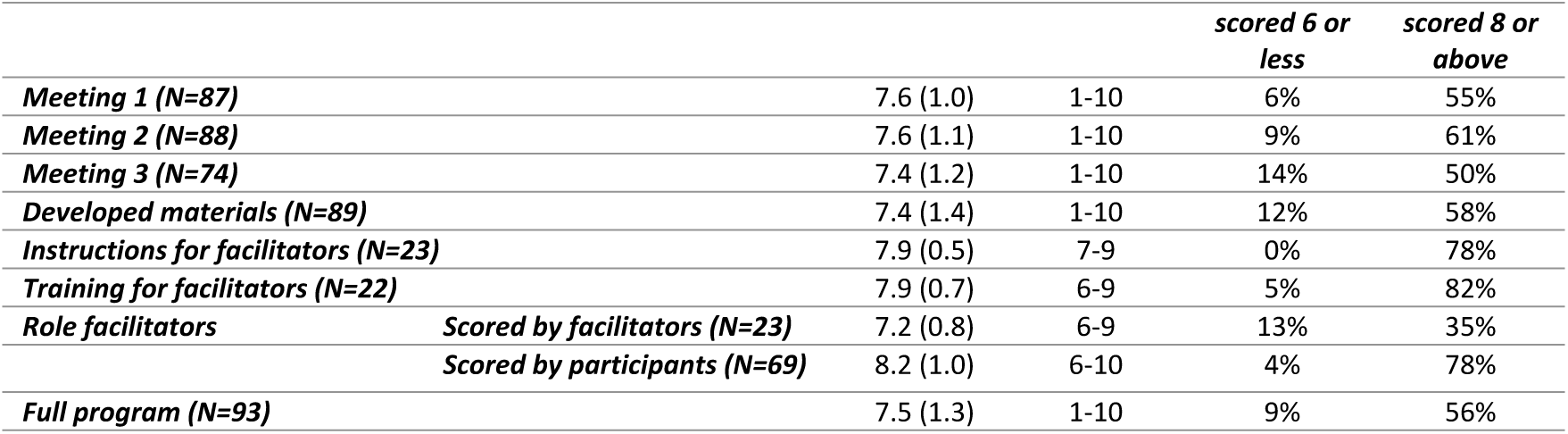
Grades for different aspects of the intervention program.

The most frequently mentioned positive aspects of the program were the awareness it raised about prevention and the interaction and discussions with colleagues. Additionally, many OPs were satisfied with the structure of the program, which repeatedly focused on prevention and helped to address preventive tasks in practice. This was confirmed during the interviews, where almost all OPs indicated that being more actively engaged with the theme of prevention was the most appealing aspect: *"To take a moment and realize that you’re already doing some preventive work. It’s also really nice to hear from each other: What are you doing? What are your experiences? How do you approach certain things? So, I found exchanging information very enjoyable. It was like getting a glimpse into colleagues’ practices. And to become more aware of it. When are you being preventive? When is it about absenteeism? What is the overlap between them? Just brainstorming with each other is always useful." [Interview 08; OP]*

Opinions on the content of the materials (e.g. instructions, training for coordinators) were mixed. A number of OPs enjoyed the creative assignments, such as the assignment to take a photo of a barrier encountered in their work, and felt that this approach could be further expanded. Others, however, found the materials too ‘school-like’, which made them less engaging. Several participants expressed a need for more guidelines and information about proven effective interventions that can be implemented, and the assignments could be made more concrete based on this. The Prevention Cycle, which was the central topic of the second meeting, was generally positively evaluated. Although the prevention cycle is an existing tool for Dutch OPs, many participants were not familiar with it. They indicated that it provided new insights and serves as a reminder to focus on prevention: *“The biggest eye-opener from that meeting was realizing that you can go through the cycle multiple times and make progress each time. Now, we even jokingly say to each other: Guys, we’ve reached the next level. This gives us the feeling that, even though it seems endless and we’re in a large, bureaucratic, political organization, we’re playing our game, completing all the levels, and advancing to the next one.” [Interview 13; group coordinator]*

#### Appropriateness

As depicted in Figure 3, 67% (N=62) of participants agreed that the program aligned well with the ambition to spend more time on prevention in practice. Additionally, 73% (N=68) agreed that the program helped to address experienced barriers and possible solutions related to the execution of preventive tasks within the group. Finally, 54% (N=50) agreed that creating a personal action plan with concrete work goals actually helped them dedicate more time to preventive tasks in practice. Several participants indicated in the questionnaire and interviews that they appreciated the embedding within existing peer coaching groups since these groups are perceived as a safe setting, provide a good opportunity for discussions with colleagues, and require little extra effort since the meetings are already scheduled. One OP noted in an interview that it is not essential to implement this within peer coaching groups; it could also be implemented in educational settings not related to peer groups.

**Figure 3:**
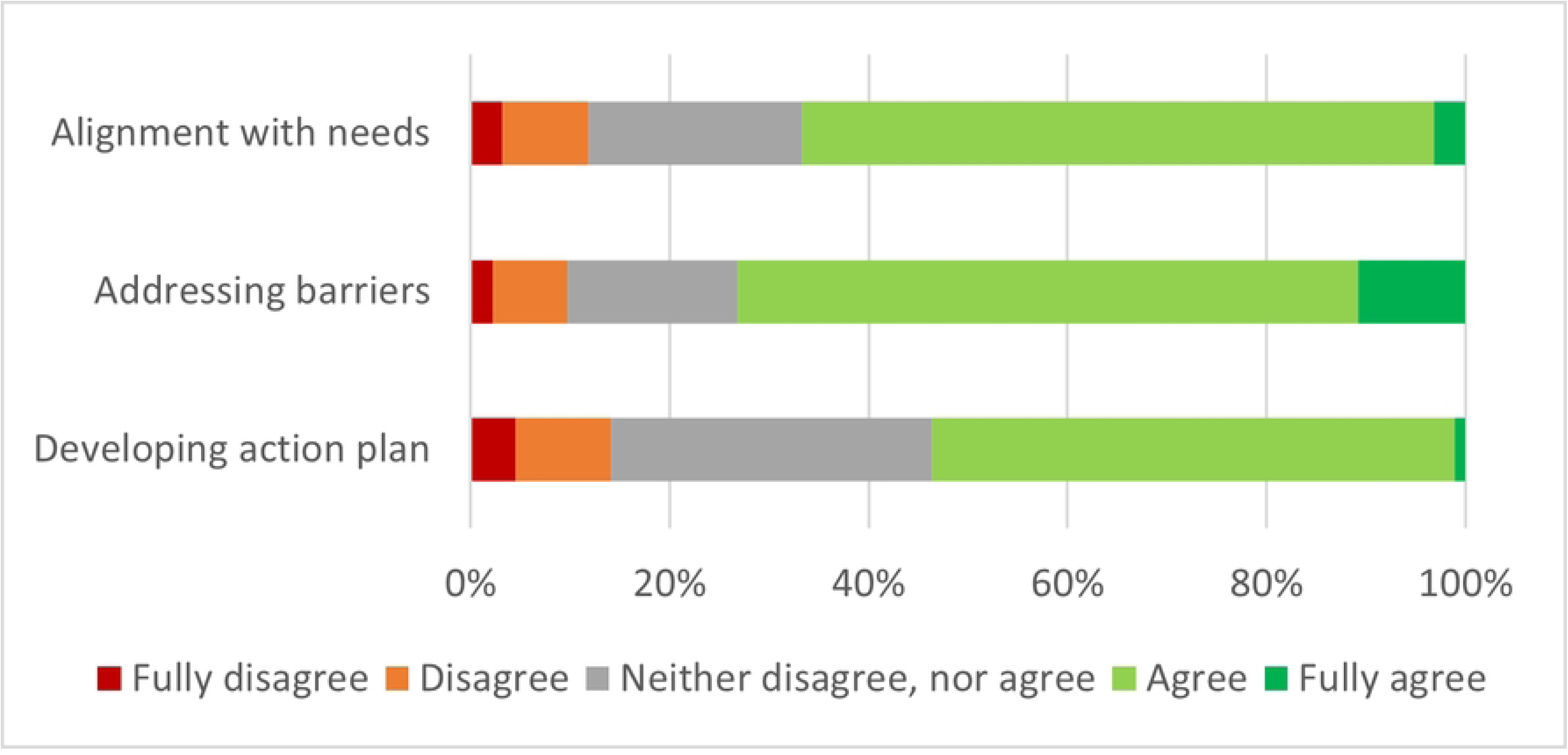
Statements on appropriateness, scored by all participants (N=93)

#### Sustainability

Several OPs indicated in the questionnaire that the program could be repeated after a certain period to maintain the focus on prevention within their work and their group. A number of participants indicated that their group had decided to already incorporate the topic of prevention into their meetings on a regular basis, for example by adding it as a fixed agenda item or dedicating one of their meetings to the topic at regular intervals: *“It remains a part of the group now, and it will be a topic at every meeting: what are we doing about prevention? How can we improve it so that we can learn from each other? That’s what we agreed on. This Thursday, we have another meeting, and it’s already on the agenda.” [Interview 10; group coordinator]*

## Discussion

### Main results

This process evaluation served four goals: 1) describe the reach and uptake of a peer coaching intervention program for OPs aimed at enhancing their preventive role; 2) examine the extent to which the program was implemented as intended; 3) identify barriers and facilitators that may have impacted the implementation; 4) and explore the experiences and perspectives of OPs, perceived fit and benefits of the intervention program.

Findings indicate that the overall program was well-implemented, increased awareness, and provided a structured approach to support OPs in integrating preventive tasks into daily practice. Participating OPs also appreciated the repeated focus on prevention, the practical tools provided, and the way the program facilitated valuable interactions among colleagues. One explanation for the positive evaluation of the program is that it meets all of the key criteria necessary for successful peer coaching among health care professionals: it needs to be voluntary, mutually beneficial, co-operative, and non-evaluative; it should focus on enhancing strengths of participants, be goal-directed and involve feedback and self-evaluation [21]. The program was implemented into the infrastructure of existing peer groups, as part of the CME system, which was also positively evaluated by OPs. The groups were ideally suited to discuss the complex issue of prevention and work towards improving the execution of their preventive tasks in practice. The use of the existing infrastructure of peer groups was not only positively evaluated by OPs, but it is also in line with principles from implementation science, because it supports integration of the intervention [22]. Moreover, using existing groups takes away some of the challenges known in group intervention implementation, such as participant recruitment and attrition [14].

Although many participants succeeded in formulating and achieving work-related goals, several barriers to achieving their set goals were mentioned by OPs, including resistance or other priorities from employers, and time constraints. These factors are also frequently cited as barriers to prevention in the scientific literature [23–25]. However, it is important to note that the extent to which OPs in this study experienced barriers was influenced by work-related factors such as sector, organization size, or type of OHS. OPs working for an in-house OHS reported having shorter lines of communication with management, making it easier for them to influence decision-making and achieve their goals, while OPs working for external OHS sometimes experienced a lack of autonomy in deciding what topics to work on. This finding aligns with a scoping review on the working conditions of OPs, which indicates that in-house positions or working as a self-employed OP are considered job resources, whereas working for an OHS can be seen as a stressor [25]. To change OPs’ work, it is important to address these barriers and power dynamics that might exist between OPs, OHS and employers. In a previous study about stimulation of the implementation of workers’ health surveillance activities, OPs were trained in having conversations with employers about the implementation of workers’ health surveillance (WHS), resulting in increased skills and self-efficacy among OPs [26]. It is therefore recommended to provide OPs with additional training in negotiation and communication skills to better advocate for other preventive measures with employers and their OHS as well, in order to take away some of the experienced barriers. In addition, it is recommended to provide employers and employees with information about the preventive role of OPs and the added value of investing in prevention, to tackle barriers such as a lack of knowledge about the value of prevention among employees or potential resistance from employers to focus on prevention [7, 9]. Moreover, according to OPs in this study, there is already significant focus on health and prevention in certain sectors in the Netherlands, such as construction and healthcare, which worked as a facilitator. In contrast, OPs working with SMEs may experience greater distance to employers and employees, making it more challenging to achieve their goals [27]. Based on these findings, it is recommended to address context-specific barriers, by tailoring strategies to the specific needs and conditions of various sectors, and to provide additional training, resources, and support specifically designed for OPs working in SMEs.

Despite the overall positive evaluation, some areas for improvement were identified. First of all, the frequency and duration of meetings could be reconsidered. While some participants found the existing schedule appropriate, others suggested a longer duration for the meetings or longer intervals between meetings to allow more time for implementing preventive tasks. A meta-analysis of studies evaluating the CME interventions concluded that longer interventions, multiple sessions or repetition over time are more effective than short or single interventions [28]. Although the current intervention meets these criteria, the time between meetings and the duration can be adjusted to the needs of each group. Second, a number of participants expressed a need for more scientific knowledge about the effectiveness of preventive interventions and how best to implement them. The intervention in this study focused on promoting the execution of preventive tasks, rather than implementing specific preventive interventions, so no conclusions can be drawn about the effect of interventions. However, within these tasks, further specific interventions can be carried out if desired. Although the body of evidence is still small, recent reviews have shown that preventive interventions and implementation strategies can have positive effects [29, 30]. New scientific knowledge should be further integrated into guidelines and tools to meet the needs of OPs and facilitate its implementation in the work of OPs. However, previous studies found that OPs may encounter barriers when using guidelines, such as knowledge or attitude-related issues and organizational barriers [31, 32]. Consistent with earlier studies, it is therefore recommended to supplement guideline development with training and practice in these areas, to facilitate learning from one another and ensure effective implementation.

### Strengths and limitations

This study has several strengths, of which the first is the mixed-methods study design, because integrating both quantitative data (questionnaire) and qualitative data (interviews) provides a more complete and nuanced understanding of the implementation process and experiences of participants. Additionally, the interviewed participants differed in aspects such as age, years of work experience, and the types of companies they work for (e.g. sector, SME or larger organizations). This diversity resulted in rich data that is expected to be applicable to many OPs. A second strength is that during the development of the questionnaire and interview guide the implementation outcomes as defined by Proctor were systematically incorporated [16]. This approach provides the advantage of offering a comprehensive view of the implementation factors and successes. Moreover, the implementation indicators focus on the engagement and needs of the target group, which provides guidance for adapting the intervention to better align with daily practice. Third, the intervention program was applied to existing peer groups and practices, requiring little to no extra time for OPs to participate in the program. This is supported by the central principle in implementation science that strategies are most successful if they align with existing infrastructures, practices and culture [22, 33]. However, each group has their own working methods, different experiences (for example due to various sectors), and individual perspectives on the topic of prevention. Even though the program and developed materials provided a clear structure, differences between groups could potentially have led to variations in implementation.

### Conclusion

A peer group coaching intervention program was developed to support OPs in their preventive role. Based on this process evaluation, it appeared that the program was well-implemented, successfully increased awareness of the relevance of prevention, contributed to valuable discussions between OPs, and helped them to formulate barriers regarding executing preventive tasks and strategies to overcome these. While a large group of OPs was able to implement their personal action plan in practice and to focus more on the prevention of work related disease, others faced resistance from employers or experienced time constraints in changing their working methods. The intervention program can be improved by including more evidence-based interventions, tools and guidelines to the materials, and allocating more time to the intervention meetings.

## Data Availability

All datafiles will be available from the data.rivm.nl database after completion of the study.

## Abbreviations

ASE: Attitude, social influence and self-efficacy
CME: Continuing Medical Education
IM: Implementation Mapping
IM-PROmPt-study: Implementation of PReventive tasks by Occupational Physicians Study
MIDI: Measurement Instrument for Determinants of Innovation
OHS: Occupational health and safety services
OP(s): Occupational physician(s)
SME: Small and Medium Enterprises
RCT: Randomized Controlled Trial
WHS: Workers ’ Health Surveillance

## Declarations

### Consent for publication

Not applicable.

### Availability of data and materials

The datasets used and/or analysed during the current study are available from the corresponding author on reasonable request.

### Authors’ contributions

SOP, SO, FS and KP were involved in the study design. SOP was involved in data collection and analyses. All authors have contributed significantly to the final manuscript and gave their approval.

## Acknowledgements

Not applicable.

## Authors’ information (optional)

Not applicable.

